# Short-term improvement of mental health after a COVID-19 vaccination

**DOI:** 10.1101/2022.02.22.22271327

**Authors:** Charilaos Chourpiliadis, Anikó Lovik, Anna K. Kähler, Unnur A. Valdimarsdóttir, Emma M. Frans, Fredrik Nyberg, Patrick F. Sullivan, Fang Fang

## Abstract

**Importance:** The role of COVID-19 vaccination on the mental health of the general population remains poorly understood.

**Objective:** To assess the short-term change of depressive and anxiety symptoms in relation to COVID-19 vaccination among Swedish adults.

**Design:** A prospective cohort study with monthly data collections on self-reported depressive and anxiety symptoms from December 2020 to October 2021 and COVID-19 vaccination from July to October 2021.

**Setting:** The Omtanke2020 Study, Sweden.

**Participants:** 7,925 participants of the Omtanke2020 study with complete data on depressive and anxiety symptoms and vaccination status.

**Intervention(s) or Exposure(s):** Receiving the first or second dose of a COVID-19 vaccine.

**Main outcomes(s) and Measure(s):** Binary measures of depression (PHQ-9, cut-off ≥10) and anxiety (GAD-7, cut-off ≥10) one month before the first dose, one month after the first dose, and, if applicable, one month after the second dose. For individuals not vaccinated or chose to not report vaccination status (unvaccinated individuals), we selected three monthly measures of PHQ-9 and GAD-7 with 2-month intervals in-between based on data availability.

**Results:** 5,079 (64.1%) individuals received two doses of COVID-19 vaccine, 1,977 (24.9%) received one dose, 305 (3.9%) were not vaccinated, and 564 (7.1%) chose not to report vaccination status. There was a lower prevalence of depression and anxiety among vaccinated, compared with unvaccinated, individuals, especially after the second dose. Among individuals receiving two doses of vaccine, the prevalence of depression and anxiety was lower after both first (aRR=0.82, 95%CI 0.76-0.88 for depression; aRR=0.81, 95%CI 0.73-0.89 for anxiety) and second (aRR=0.79, 95%CI 0.73-0.85 for depression; aRR=0.73, 95%CI 0.66-0.81 for anxiety) dose, compared with before vaccination. Similar results were observed among individuals receiving only one dose (aRR=0.76, 95%CI 0.68-0.84 for depression; aRR=0.82, 95%CI 0.72-0.94 for anxiety, comparing after first dose to before vaccination). These results were independent of age, sex, recruitment type, body mass index, smoking, relationship status, history of psychiatric disorder, number of comorbidities, COVID-19 infection status, and seasonality.

**Conclusions and Relevance:** We observed a positive short-term change in depressive and anxiety symptoms among adults receiving a COVID-19 vaccine in the current pandemic.

**Key points:** *Question:* Is COVID-19 vaccination associated with a short-term change in mental health?

*Findings:* This longitudinal study included 7,925 Swedish adults with self-reported COVID-19 vaccination and symptoms of mental health responding from December 2020 to October 2021. The prevalence of depressive or anxiety symptoms was lower one month after vaccination compared to one month before vaccination. The effect size was greater among individuals receiving two doses of vaccine, compared with those receiving only one dose.

*Meaning:* Receiving vaccination against COVID-19 is associated with short-term improvement in mental health.

## Introduction

The COVID-19 pandemic has caused an unprecedented health crisis worldwide. The negative mental health impact of the pandemic has been demonstrated, affecting both the infected and non-infected individuals (1, 2). Vaccination against COVID-19 has been suggested to have a beneficial impact on mental health, in addition to protection from severe infection (3). However, the existing evidence rests on a restricted phase of the vaccine rollout (4) and studies with relatively small sample size (5).

Our aim was to provide an estimate of the short-term change in mental health after the vaccination against COVID-19, specifically depression and anxiety, during a long phase of the vaccine rollout and using a large study sample with careful adjustment for the impact of age, sex, history of COVID-19 infection, etc. We hypothesized that receiving a COVID-19 vaccine was associated with a decrease in the prevalence of depressive and anxiety symptoms.

## Methods

We performed a longitudinal study based on the Omtanke2020 study, an ongoing prospective cohort study specifically designed to assess the mental health impact of COVID-19 in Sweden (6). Over 28,000 adult Swedish residents were recruited to Omtanke2020 between June 2020 and June 2021 and provided self-reported data on mental and physical health, sociodemographic factors, lifestyle factors, and COVID-19-specific questions such as infection and vaccination (6, 7).

In the present study, we included all participants of Omtanke2020 with monthly data on depressive and anxiety symptoms between December 2020 and October 2021. These participants also had to have data on COVID-19 vaccination collected between 17^th^ July and 30^th^ October 2021. We excluded from the analysis 40 participants with inconsistent information on vaccination (e.g., time of the 2^nd^ dose preceding that of the 1^st^ dose) and 22 participants who received the 1-dose Janssen vaccine. We defined the vaccinated individuals as those reporting having received two doses or one dose of a COVID-19 vaccine, and the unvaccinated individuals as those reporting no vaccination or chose not to report vaccination status.

The Patient Health Questionnaire (PHQ-9) (8) was used to measure depressive symptoms whereas the Generalized Anxiety Disorder (GAD-7) (9) scale was used to measure anxiety symptoms. The binary outcomes of depression and anxiety were calculated from the total scores of these scales, which are shown to have appropriate psychometric properties in Omtanke2020 (6). A cut-off of 10 in each scale was used to define significant depressive or anxiety symptoms, respectively (8, 9).

For vaccinated individuals, we considered depression and anxiety at three timepoints: one month before 1^st^ dose (baseline), first month after the first dose, and, if applicable, first month after the second dose. For unvaccinated individuals, we randomly selected three timepoints with a 2-month interval in-between during the study period (i.e., baseline/Time 0, 2 months after baseline/Time 1, and 4 months after baseline/Time 2).

We first used Poisson regression to calculate prevalence ratio, comparing the prevalence of depression and anxiety between vaccinated and unvaccinated individuals, at baseline, after 1^st^ dose/Time 1, and after 2^nd^ dose/Time 2. To disentangle the seasonality effect from the vaccine effect, we performed a stratified analysis by calendar month. We first calculated crude prevalence ratio and then adjusted the analysis for age, sex, body mass index (BMI), recruitment type (self-recruitment or by invitation), relationship status, current smoking, number of comorbidities, history of psychiatric disorder, history of COVID-19 infection, and month of baseline survey.

We also used a Generalized Estimating Equations (GEE) model to assess the temporal change in the prevalence of depression and anxiety among vaccinated and unvaccinated individuals separately. In this analysis, we adjusted for age, sex, BMI, recruitment type, relationship status, current smoking, number of comorbidities, history of psychiatric disorder, history of COVID-19 infection, and month of baseline survey and used robust standard error to correct for the overestimated standard error of a log-Poisson model with binary outcome (10).

## Results

A total of 7,925 individuals, with a mean age of 52.6 (±15.4) years and 83% women, were included in the analysis, including 5,079 (64.09%) with two doses and 1,977 (24.95%) with one dose of COVID-19 vaccine, 305 (3.85%) not vaccinated, and 564 (7.12%) not reporting vaccination status (Table 1).

**Table 1.**
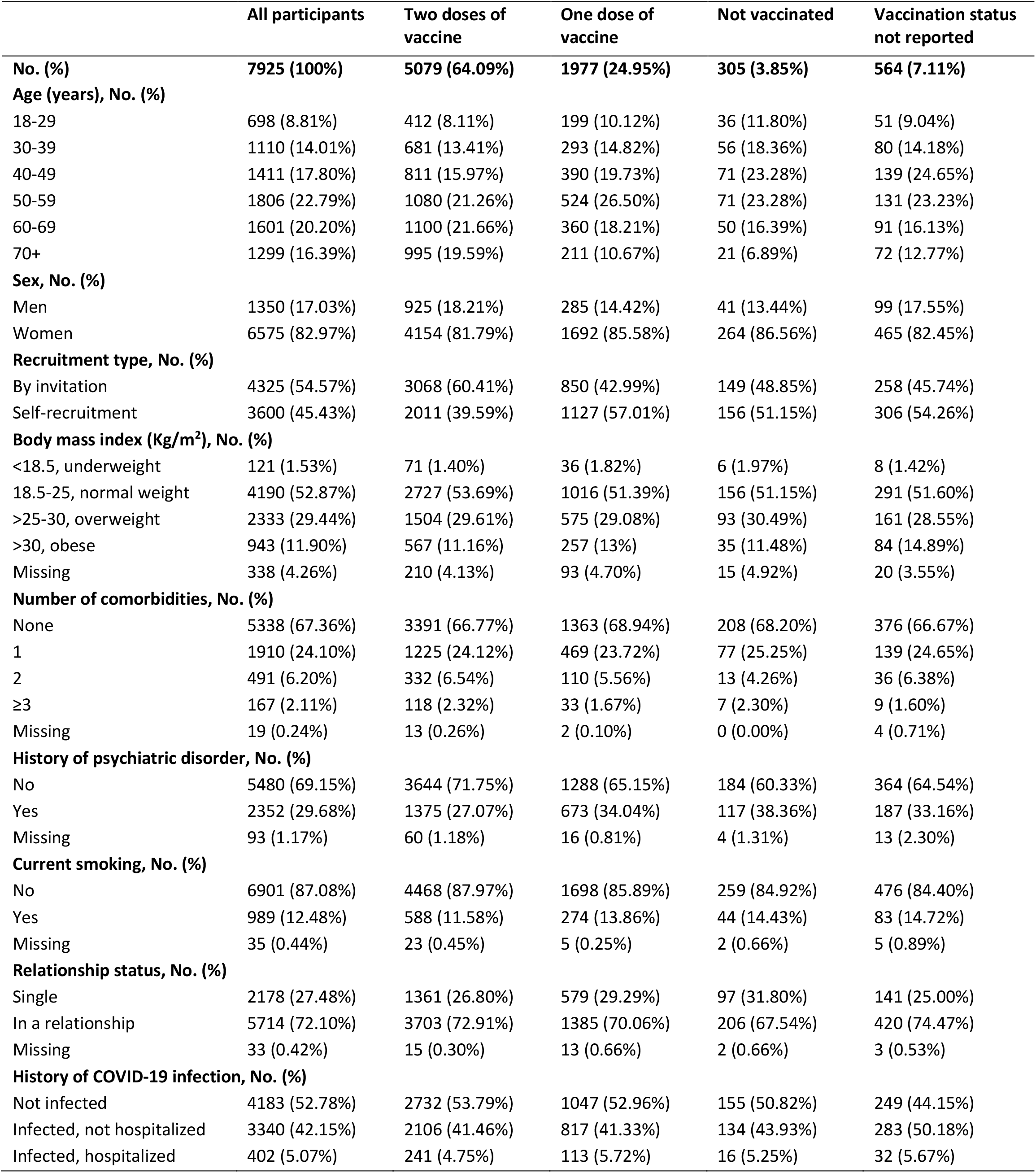
Baseline characteristics of the study participants by vaccination status.

**Table 2.**
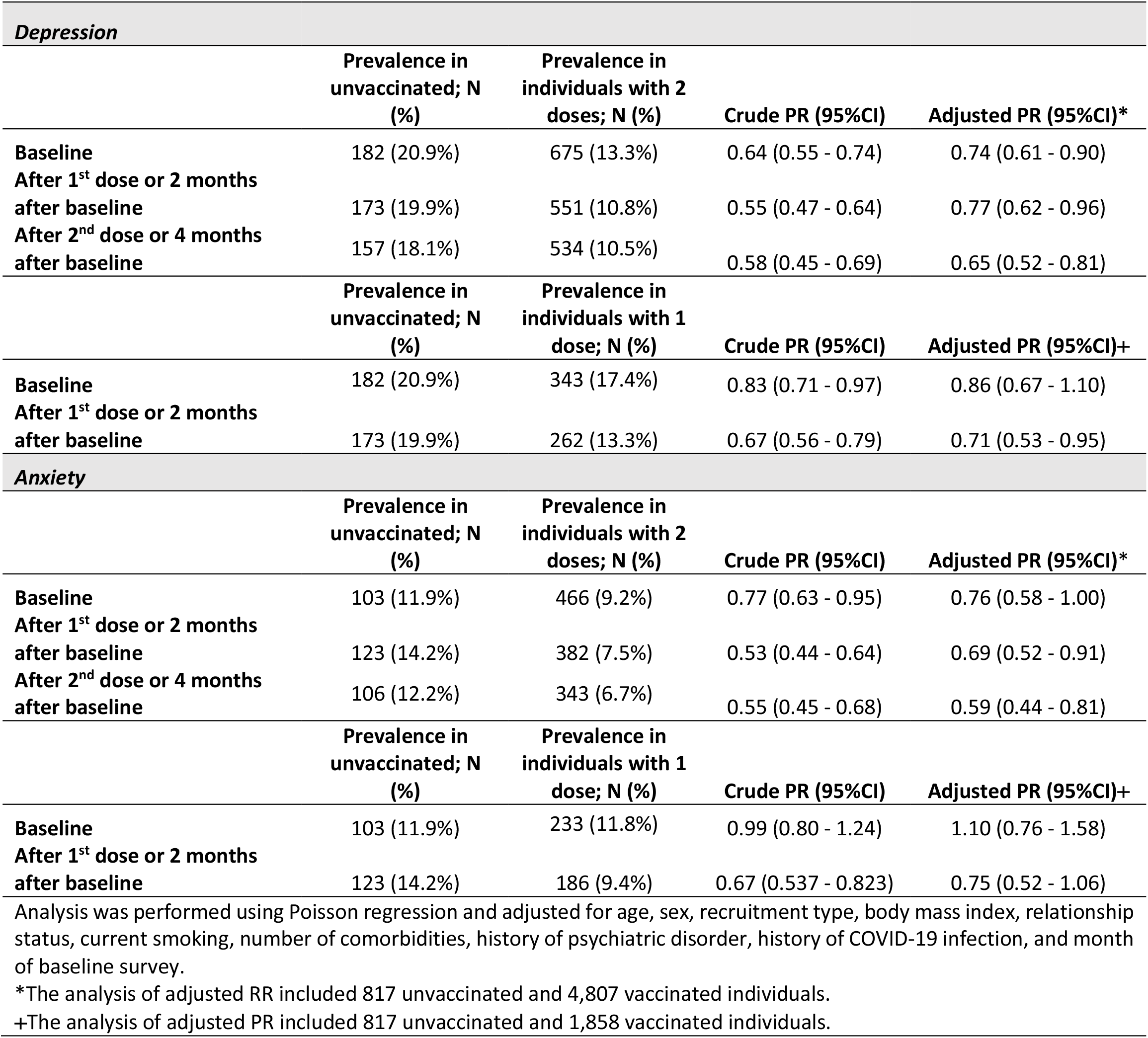
Prevalence ratio (PR) and 95% confidence interval (CI) of depression and anxiety comparing vaccinated to unvaccinated individuals.

| *Depression* |  |  |  |  |
| --- | --- | --- | --- | --- |
|  | Prevalence in unvaccinated; N (%) | Prevalence in individuals with 2 doses; N (%) | Crude PR (95%CI) | Adjusted PR (95%CI)* |
| Baseline | 182 (20.9%) | 675 (13.3%) | 0.64 (0.55 - 0.74) | 0.74 (0.61 - 0.90) |
| After 1<sup>st</sup> dose or 2 months after baseline | 173 (19.9%) | 551 (10.8%) | 0.55 (0.47 - 0.64) | 0.77 (0.62 - 0.96) |
| After 2<sup>nd</sup> dose or 4 months after baseline | 157 (18.1%) | 534 (10.5%) | 0.58 (0.45 - 0.69) | 0.65 (0.52 - 0.81) |
|  | Prevalence in unvaccinated; N (%) | Prevalence in individuals with 1 dose; N (%) | Crude PR (95%CI) | Adjusted PR (95%CI)+ |
| Baseline | 182 (20.9%) | 343 (17.4%) | 0.83 (0.71 - 0.97) | 0.86 (0.67 - 1.10) |
| After 1<sup>st</sup> dose or 2 months after baseline | 173 (19.9%) | 262 (13.3%) | 0.67 (0.56 - 0.79) | 0.71 (0.53 - 0.95) |
| *Anxiety* |  |  |  |  |
|  | Prevalence in unvaccinated; N (%) | Prevalence in individuals with 2 doses; N (%) | Crude PR (95%CI) | Adjusted PR (95%CI)* |
| Baseline | 103 (11.9%) | 466 (9.2%) | 0.77 (0.63 - 0.95) | 0.76 (0.58 - 1.00) |
| After 1<sup>st</sup> dose or 2 months after baseline | 123 (14.2%) | 382 (7.5%) | 0.53 (0.44 - 0.64) | 0.69 (0.52 - 0.91) |
| After 2<sup>nd</sup> dose or 4 months after baseline | 106 (12.2%) | 343 (6.7%) | 0.55 (0.45 - 0.68) | 0.59 (0.44 - 0.81) |
|  | Prevalence in unvaccinated; N (%) | Prevalence in individuals with 1 dose; N (%) | Crude PR (95%CI) | Adjusted PR (95%CI)+ |
| Baseline | 103 (11.9%) | 233 (11.8%) | 0.99 (0.80 - 1.24) | 1.10 (0.76 - 1.58) |
| After 1<sup>st</sup> dose or 2 months after baseline | 123 (14.2%) | 186 (9.4%) | 0.67 (0.537 - 0.823) | 0.75 (0.52 - 1.06) |

Analysis was performed using Poisson regression and adjusted for age, sex, recruitment type, body mass index, relationship status, current smoking, number of comorbidities, history of psychiatric disorder, history of COVID-19 infection, and month of baseline survey.
\*The analysis of adjusted RR included 817 unvaccinated and 4,807 vaccinated individuals.
+The analysis of adjusted PR included 817 unvaccinated and 1,858 vaccinated individuals.

Although there tended to be a lower prevalence of depression and anxiety among the vaccinated individuals, compared with the unvaccinated individuals, already at baseline, the difference became greater after the 2^nd^ dose among those with two doses of vaccine and after the 1^st^ dose among those with one dose of vaccine (Table 1). The stratified analysis by calendar month showed a similar result pattern across different calendar months (Supplementary Table 1).

Figure 1 shows the risk ratio (RR) of depression and anxiety, comparing the prevalence of depression and anxiety after a COVID-19 vaccine (1^st^ or 2^nd^ dose) to the prevalence before vaccination or the prevalence at later time points (Time 1 or 2) to the prevalence at first time point (Time 0) among the unvaccinated individuals. There was a decline in the prevalence of depression and anxiety among individuals that received two doses of vaccine, both during the month after 1^st^ dose (aRR=0.82; 95%CI: 0.76-0.88 for depression; aRR=0.81; 95%CI: 0.73-0.89 for anxiety) and during the month after 2^nd^ dose (aRR=0.79; 95%CI: 0.73-0.85 for depression; aRR=0.73; 95%CI: 0.66-0.81 for anxiety). A decrease in prevalence was also noted during the month after the 1^st^ dose among individuals that received only one dose of vaccine (aRR=0.76; 95CI: 0.68-0.84 for depression; aRR=0.82; 95%CI: 0.72-0.94 for anxiety). Among the unvaccinated individuals, there was no statistically significant decrease in the prevalence of depression or anxiety over time, except when comparing the prevalence of depression at Time 2 to that of Time 0.

**Figure 1.**
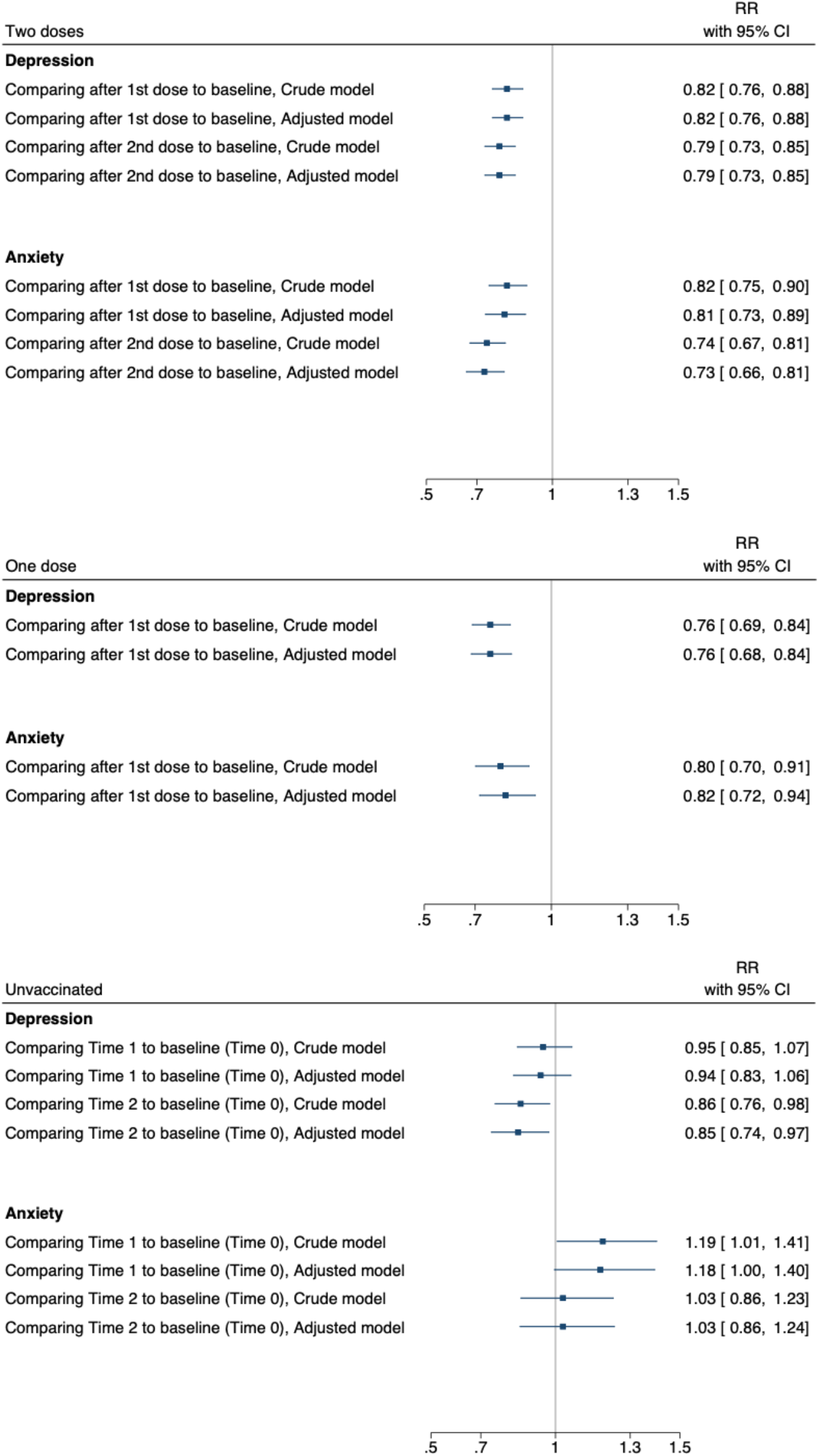
Risk Ratio (RR) of depression and anxiety, comparing the prevalence of depression and anxiety after COVID-19 vaccine (1^st^ or 2^nd^ dose) to the prevalence before vaccine among vaccinated individuals, and comparing the prevalence of later time points to baseline among the unvaccinated individuals. Unvaccinated individuals include individuals reporting no vaccination or chose to not report vaccination status. The analyses were performed using GEE models and adjusted for age, sex, recruitment type, body mass index, relationship status, current smoking, number of comorbidities, history of psychiatric disorder, history of COVID-19 infection, and month of baseline survey

## Discussion

The present study demonstrated a short-term improvement in the prevalence of depression and anxiety after COVID-19 vaccination in a large sample of Swedish adults. The improvement was more pronounced after the second dose, compared with the first dose, and was independent of age, sex, history of COVID-19 infection, etc.

Our results are consistent with previous research suggesting a positive influence of COVID-19 vaccination on mental health (3-5, 11). The present study extends however the existing knowledge by addressing several methodological issues of previous studies, such as cross-sectional study design (5, 11), relatively small sample size (5), the use of a single mental health instrument (e.g., PHQ-4) (3, 4, 11), and addressing a restricted phase of vaccination, e.g., March 2021 (3) or June 2021 (4). First, our study used a longitudinal design with monthly surveys on vaccination status and mental health which allowed us to understand the effect of COVID-19 vaccine on the temporal change of depression and anxiety as well as to disentangle the effect of vaccine from seasonal variation in the prevalence of depression and anxiety. The large sample size, the study of both depression and anxiety using well-established instruments, and the relatively long duration of the study period are other strengths. Limitations of the study include self-reported data on COVID-19 vaccination and depressive and anxiety symptoms. Finally, whether our results are generalizable to other populations remains to be studied due to the largely different burden of pandemic and population coverage of COVID-19 vaccination across the globe.

## Conclusion

We found an immediate positive change in symptoms of depression and anxiety after receiving a COVID-19 vaccination among adults in the current pandemic.

## Data Availability

All data produced in the present study are available for a specified purpose after approval by the institution and the principal investigator of the Omtanke2020 study and with a signed data access agreement.

## Acknowledgement

This study was supported by NordForsk (project No. 105668) and Horizon2020 (CoMorMent, 847776). The authors also thank the participants of Omtanke2020.

## Ethical approval

The study was approved (DNR 2020-01785) by the Swedish Ethical Review Authority on 3 June 2020.

