## Supplemental Table 1 for "Short-term improvement of mental health after a COVID-19 vaccination"

| **Supplementary eTable 1. Prevalence (95% confidence interval) of depression and anxiety among vaccinated and unvaccinated individuals, by calendar month in 2021.** | | | | | | | | | | | | | | |
| --- | --- | --- | --- | --- | --- | --- | --- | --- | --- | --- | --- | --- | --- | --- |
| ***Depression (crude prevalence)*** | | | | | | | | | | | | | | |
|  | February | March | | April | | May | | June | July | August | September | | October | |
| **Unvaccinated** (N) | 6330 | 6606 | | **4647** | | **3146** | | **1575** | **514** | **269** | 185 | | 127 | |
| **Unvaccinated** | 0.16 (0.15 - 0.17) | 0.16 (0.15 -0.17) | | **0.18 (0.17 - 0.19)** | | **0.19 (0.17 - 0.20)** | | **0.20 (0.18 - 0.22)** | **0.17 (0.14 - 0.20)** | **0.17 (0.12 - 0.21)** | 0.17 (0.12 - 0.23) | | 0.17 (0.10 - 0.23) | |
| **Vaccinated** (N) | 97 | 705 | | **2574** | | **4413** | | **5744** | **6489** | **5929** | 4273 | | 3136 | |
| **Vaccinated** | 0.20 (0.12 - 0.28) | 0.13 (0.10 - 0.15) | | **0.10 (0.09 - 0.12)** | | **0.11 (0.10 - 0.12)** | | **0.11 (0.10 - 0.12)** | **0.10 (0.09 - 0.11)** | **0.10 (0.09 - 0.11)** | 0.12 (0.11 - 0.13) | | 0.13 (0.11 - 0.14) | |
| ***Depression (adjusted prevalence)*** | | | | | | | | | | | | | | |
| **Unvaccinated** (N) | 5972 | 6237 | 4367 | | 2932 | | 1433 | | 478 | 253 | | 170 | | 121 |
| **Unvaccinated** | 0.15 (0.14 - 0.16) | 0.15 (0.14 - 0.16) | 0.15 (0.14 - 0.16) | | 0.14 (0.13 - 0.15) | | 0.14 (0.12 - 0.15) | | 0.13 (0.11 - 0.16) | 0.15 (0.11 - 0.19) | | 0.15 (0.10 - 0.20) | | 0.16 (0.10 - 0.22) |
| **Vaccinated** (N) | 93 | 671 | 2451 | | 4214 | | 5471 | | 6131 | 5600 | | 4034 | | 2957 |
| **Vaccinated** | 0.15 (0.09 - 0.21) | 0.15 (0.12 - 0.17) | 0.14 (0.12 - 0.15) | | 0.14 (0.12 - 0.15) | | 0.12 (0.11 - 0.13) | | 0.10 (0.09 - 0.11) | 0.10 (0.09 - 0.11) | | 0.12 (0.11 - 0.13) | | 0.12 (0.11 - 0.13) |
| ***Anxiety (crude prevalence)*** | | | | | | | | | | | | | | |
| **Unvaccinated** (N) | 6330 | 6606 | **4647** | | **3146** | | **1575** | | **514** | 269 | | **185** | | 127 |
| **Unvaccinated** | 0.11 (0.10 - 0.12) | 0.11 (0.10 - 0.12) | **0.12 (0.11 - 0.13)** | | **0.14 (0.12 - 0.15)** | | **0.15 (0.14 - 0.17)** | | **0.12 (0.09 - 0.15)** | 0.11 (0.07 - 0.15) | | **0.15 (0.10 - 0.20)** | | 0.11 (0.06 - 0.17) |
| **Vaccinated** (N) | 97 | 705 | **2574** | | **4413** | | **5744** | | **6489** | 5929 | | **4273** | | 3136 |
| **Vaccinated** | 0.14 (0.07 - 0.2) | 0.08 (0.06 - 0.10) | **0.06 (0.05 - 0.07)** | | **0.07 (0.06 - 0.08)** | | **0.08 (0.07 - 0.08)** | | **0.07 (0.06 - 0.07)** | 0.07 (0.07 - 0.08) | | **0.08 (0.08 - 0.09)** | | 0.10 (0.09 - 0.11) |
| ***Anxiety (adjusted prevalence)*** | | | | | | | | | | | | | | |
| **Unvaccinated** (N) | 5972 | 6237 | 4367 | | 2932 | | 1433 | | 478 | 253 | | 170 | | 121 |
| **Unvaccinated** | 0.11 (0.10 - 0.11) | 0.10 (0.10 - 0.11) | 0.10 (0.09 - 0.11) | | 0.10 (0.09 - 0.11) | | 0.10 (0.09 - 0.12) | | 0.09 (0.07 - 0.11) | 0.09 (0.06 - 0.12) | | 0.13 (0.08 - 0.18) | | 0.10 (0.05 - 0.16) |
| **Vaccinated** (N) | 93 | 671 | 2451 | | 4214 | | 5471 | | 6131 | 5600 | | 4034 | | 2957 |
| **Vaccinated** | 0.12 (0.07 - 0.18) | 0.10 (0.07 - 0.12) | 0.09 (0.07 - 0.11) | | 0.09 (0.08 - 0.10) | | 0.08 (0.08 - 0.09) | | 0.07 (0.06 - 0.07) | 0.07 (0.06 - 0.08) | | 0.08 (0.07 - 0.09) | | 0.10 (0.09 - 0.11) |
| Adjusted prevalence was adjusted for age, sex, recruitment type, body mass index, relationship status, current smoking, number of comorbidities, history of psychiatric disorder, and history of COVID-19 infection. **Bold** font indicates statistically significant difference between vaccinated and unvaccinated individuals. | | | | | | | | | | | | | | |
